# Orbitofrontal thickness and network associations as transdiagnostic signature of negative symptoms along the bipolar-schizophrenia spectrum

**DOI:** 10.1101/2024.07.29.24311172

**Authors:** Marlene Franz, Valeria Kebets, Xaver Berg, Foivos Georgiadis, Achim Burrer, Janis Brakowski, Stefan Kaiser, Erich Seifritz, Philipp Homan, Esther Walton, Theo G. M. van Erp, Jessica A. Turner, Bratislav Misic, Sofie L. Valk, B.T. Thomas Yeo, Boris C. Bernhardt, Matthias Kirschner

## Abstract

Negative symptoms are core features of schizophrenia (SCZ) and also prevalent in bipolar disorder (BD). While orbitofrontal cortex (OFC) alterations have been implicated in the development of negative symptoms, their contributions across disorders remain to be established. Here, we tested how OFC thickness and related network associations relate to severity of negative symptom dimensions across the BD-SCZ spectrum. We included 50 individuals with SCZ, 49 with BD, alongside 122 controls. We assessed amotivation and diminished expression and estimated thickness in the medial and lateral OFC as regions-of-interest as well as 64 other cortical regions. Across BD and SCZ, reduced right lateral and bilateral medial OFC thickness were specifically associated with amotivation, but not diminished expression or other clinical factors. We then generated OFC structural co-variation networks to evaluate how the system-level embedding of the OFC would link to brain-wide cortical maps of negative symptoms. We found that medial OFC co-variation networks spatially correlated with the cortical maps of both negative symptom dimensions. Confirmatory analyses in independent SCZ data from the ENIGMA consortium (n=4,474) revealed similar associations with lateral OFC co-variation networks. Finally, the brain-wide cortical alteration pattern of amotivation was significantly correlated with normative functional and structural white-matter connectivity profiles of the right medial and left lateral OFC as well as adjacent prefrontal and limbic regions. Our work identifies OFC alterations as a possible transdiagnostic signature of amotivation and provide insights into network associations underlying the system-wide cortical alterations of negative symptoms across SCZ and BD.

## Introduction

Negative symptoms are a core feature of schizophrenia (SCZ) and greatly impact the clinical outcome and quality of life in affected individuals.^1–4^ The current conceptualization of negative symptoms comprises five domains including avolition, anhedonia, asociality, blunted affect, and alogia.^5,6^ Psychometric studies have shown that these five domains map onto at least two dimensions. Avolition, anhedonia and asociality cluster within a negative motivational symptom dimension, herein defined as amotivation dimension, while alogia and blunted affect can be grouped into a diminished expression dimension.^7–10^ The amotivation dimension is characterized by a reduction in goal-directed activities, diminished interest, and lack of experiencing pleasure in current or future activities. The diminished expression dimension can be defined by a decrease in expression of spontaneous and elicited emotions and a quantitative reduction of speech. Consistent with this psychometric and phenomenological distinction, both negative symptom dimensions differentially impact long-term outcome. In this regard, in particular the amotivation dimension has been identified as strong predictor of poor functioning and diminished quality of life.^1,2,11,12^

Negative symptoms have long been thought to be specific to SCZ but have recently gained interest as transdiagnostic construct across the bipolar disorder (BD) - SCZ spectrum.^13,14^ Studies have reported comparable levels of avolition and anhedonia in SCZ and BD patients supporting transdiagnostic continuity of the amotivation dimension.^14–16^ In addition, amotivational symptoms have been related to neurocognitive impairments in both SCZ and BD,^17,18^ thus raising the possibility of shared underlying neural mechanisms. On a neural level, amotivation may reflect prefrontal-striatal reward network alterations.^19–21^ Functional resonance imaging (fMRI) studies in SCZ consistently relate higher amotivation severity to altered prefrontal-striatal reward signals and connectivity.^22–30^ In line with this prefrontal-striatal dysfunction hypothesis, neuroanatomical alterations within these regions have also been associated with higher amotivation across neurological disorders and SCZ.^31,32^ Orbitofrontal cortex (OFC) thickness alterations have emerged as a robust neural signature of negative symptoms, particularly amotivation, across disease stages in SCZ.^33–36^ For example, a meta-analysis from the ENIGMA consortium found that reduced lateral and medial OFC thickness are correlated with higher negative symptoms in SCZ.^37^ Recent work from our group extended these findings showing that these associations are also detectable in individuals with first episode psychosis.^34^

Although amotivational negative symptoms reflect a common symptom dimension in BD and SCZ, studies examining the role of OFC as a transdiagnostic signature of this association are lacking. Investigations into the brain signatures of common symptom dimensions across BD and SCZ are further supported by evidence of overlapping morphometric alterations associated with these disorders. Recent meta-analysis and cross-disorder studies have identified shared OFC thickness reductions with comparable effect sizes.^38–40^ It has been shown that the observed patterns of cortical abnormalities reflect co-variation networks, such that regions exhibiting more similar cortical thickness variations tend to be more strongly connected to each other.^41,42^ In this regard, recent work identified associations between regional structural covariance networks and distinct symptom dimensions in BD and SCZ.^43,44^ These results support the hypothesis that brain-wide symptom-structure relationships might be guided by localized regions, which in turn have strong associations with the respective symptom. In addition, it has been shown that disease-specific cortical alterations of BD and SCZ are constrained by overlapping normative functional and structural connectome features.^45,46^ This comparison of cortical alterations to normative regional functional and structural connectivity profiles, defined as epicenter mapping, has been initially developed to identify epicenters of atrophy spread in neurodegenerative disorders.^47–49^ More recently this epicenter mapping has been validated in several studies examining disease-specific and cross-disorder epicenters in neuropsychiatric disorders. These findings support the hypothesis that disease- or symptom-specific cortical alteration patterns may propagate from distinct epicenters to other cortical regions via network mechanisms.^41,45,50^ Together these findings suggest that connectivity profile of specific brain regions shapes the disease or symptom-specific spatial pattern of cortical alterations. Based on this complementary research on the network characteristics of cortical alterations in BD and SCZ, two research questions arise. First, is the system-level embedding of OFC, as measured by OFC structural co-variation networks, associated with the whole-brain cortical alteration pattern of amotivation severity? Second, do functional or structural cortico-cortical connectivity profiles of the OFC or adjacent regions spatially correlate with the cortical alteration pattern associated with amotivation severity, indicating network propagation from putative epicenters?

Integrating these multiple lines of research, our study aims to investigate whether localized OFC thickness alterations and OFC co-variation networks constitute a transdiagnostic signature of amotivation across the BD-SCZ spectrum. We leveraged clinical and structural imaging data of individuals with BD and SCZ to test the following two hypotheses: 1) Based on evidence in SCZ, we hypothesized that reduced lateral and medial OFC thickness correlate with higher negative symptoms load across the BD-SCZ spectrum. 2) We further hypothesized that these associations are stronger for the amotivation rather than diminished expression dimension, and are not driven by other symptoms or medication dose. In exploratory follow-up analyses, we also investigated whether OFC co-variation networks are associated with the cortical effect size map related to amotivation severity across the BD-SCZ spectrum. Finally, we tested how the whole-brain amotivation related cortical alteration pattern is constrained by normative functional and structural connectivity of the OFC and other cortical regions.

## Methods

### Participants

Clinical and structural imaging data were obtained from the UCLA CNP cohort (University of California, Consortium for Neuropsychiatric Phenomics) and downloaded from the public database OpenfMRI (https://openfmri.org/dataset/ds000030/). Data from 50 patients diagnosed with SCZ and 49 patients diagnosed with BD type I were included in the main analysis of this study. Additionally, 122 healthy controls (HC) from the same UCLA CNP cohort were included for additional group comparisons (for details see supplementary methods and results). All participants were part of a larger multimodal imaging study which have been described in detail elsewhere.^51^ In brief, all participants were recruited from the Los Angeles area and included if the following inclusion criteria were met: age 21 to 50 years; primary language either English or Spanish; at least 8 years of formal education; no significant medical illness; negative drug test. HC participants were excluded if they had a lifetime diagnosis of a psychiatric disorder, substance abuse or dependence. BD type I participants were not allowed to have a diagnosis of SCZ and vice versa. All participants with BD type I or SCZ were outpatient individuals presenting clinical stability to complete the clinical assessment and extensive multimodal imaging sessions. Diagnoses were based on DSM-IV criteria using a semi-structured assessment with Structured Clinical Interview (SCID) for the DSM-IV. For details see.^51^

### Image acquisition and processing

T1-weighted structural brain scans were processed using FreeSurfer Version 5.3.0^52^ and were derived from a previous resting-state fMRI publication on the same dataset.^53^ We extracted cortical thickness for 68 Desikan-Killiany (DK) atlas regions^54^ (right and left hemisphere; 34 regions per hemisphere) and performed Quality Control (QC) using standard ENIGMA protocol (http://enigma.ini.usc.edu/protocols/imaging-protocols).^40^ After QC, four participants (HC=3, SCZ=1) were excluded due to poor segmentation resulting in a final sample of 49 patients with SCZ, 49 patients with BD and 119 controls. To account for scanner and site effects data were harmonized using ComBat.^55^ Finally, the cortical thickness of each region was corrected for age and sex and the standardized residuals were used for all subsequent analyses. To address our *a priori* hypotheses, we applied a region of interest (ROI) approach focusing on the standardized residuals of left and right lateral OFC and medial OFC thickness (total ROIs =4) (Table S1).

### Assessment of negative symptoms and other clinical factors

Negative symptom severity was assessed with the Scale for the Assessment of Negative Symptoms^56^ (SANS) using the global scores of the negative symptom domains blunted affect, alogia, avolition-apathy, and anhedonia-asociality. To measure the two negative symptom dimensions of amotivation and diminished expression, we followed the approach of previous studies, including our own work on the relationship between cortico-subcortical alterations and negative symptoms across stages of the schizophrenia spectrum.^4,34^ The amotivation dimensions was defined by the sum of the avolition-apathy and anhedonia-asociality global scores, and the diminished expression dimension by the sum of the blunted affect and alogia global scores.

In addition to negative symptoms, positive, depressive, and disorganized symptoms as well as risperidone equivalent dose (mg/d) were calculated as potential secondary sources of negative symptoms.^22,57^ To assess the severity of positive symptoms the Scale for the Assessment of Positive Symptoms (SAPS) was used.^56^ The SAPS includes the four positive symptom domains delusion, hallucination, bizarre behavior and positive formal thought. Following previous work^58,59^ the delusion and hallucination domains were combined to form the SAPS positive symptoms dimension, while the bizarre behavior and positive formal thought domains were combined to form the SAPS disorganization dimension. Depressive symptoms were assessed using the Hamilton depression scale with 21-items (HAMD 21).^60^ Risperidone equivalent dose was determined by converting doses of individual antipsychotics using the defined daily doses (DDD) method from.^61^

### Statistical analysis

Analyses were performed using SPSS Version 28.0.1.1(14) (IBM SPSS Inc.), Matlab Version 9.12.0 (R2022a), and R (2009-2021 RStudio, PBS; 2021,09.0 Build 351).

### Group comparison of OFC thickness

Prior to our main analyses of assessing the relationship between OFC thickness and amotivation across BD and SCZ, we compared the left/right lateral and medial OFC residuals (n=4)) between HC, BD, and SCZ. Using separate analysis of variance (ANOVA), we did not find any differences in OFC thickness between BD and SCZ. For details see Tables S2.

### Relationship between OFC thickness and amotivation

Spearman rank correlations were used to investigate the associations between SANS amotivation scores and the residuals of left/right lateral and medial OFC across all individuals with SCZ and BD. To validate the stability of these associations, we applied bootstrap resampling and generated 1000 samples with replacement of the bilateral lateral and medial OFC variables. We then calculated the 95% confidence interval by correlating the original SANS amotivation scores with each of the derived bootstrap samples. We further aimed to establish whether associations with lateral and medial OFC thickness reflect specific neural substrates of amotivation or are also related to the diminished expression dimension. We repeated the correlation analyses with the diminished expression dimension and compared differences in the magnitude of the correlation coefficient of the different negative symptom dimensions using Hittner et al.’s method for nonparametric correlations.^62,63^ All correlations between the four OFC ROIs and the two negative symptom dimensions were corrected for multiple comparisons using a false discovery rate (FDR) of q <0.05. Separate confirmatory correlation analyses in SCZ and BD tested whether transdiagnostic relationships between amotivation and OFC thickness were also significant in each diagnostic group.

### Relationship between OFC thickness and other clinical factors

We next assessed whether associations between OFC thickness and symptom severity are specific signatures of negative symptoms or are also influenced by other symptom dimensions or medication status. We performed exploratory spearman rank correlations between the left/right lateral OFC and medial OFC thickness with the SAPS positive symptom and SAPS disorganization dimensions, depressive symptoms (HAMD 21) and risperidone equivalent dose (mg/d).

### OFC co-variation networks and cortical alteration of amotivation

Morphological characteristics co-vary across cortical regions and form distinct cortico-cortical co-variation networks.^41,64–66^ Building on these findings, we hypothesized that cortical regions with higher degree of co-variation with OFC thickness would also be more strongly associated with amotivation. We estimated the joint cortical co-variation of each cortical region with each OFC ROI resulting in four separate OFC co-variation networks. We applied the structural covariance approach from Yun and colleagues that calculates the inverse exponential of the difference between brain morphological values.^67^ First, age and sex were regressed out from cortical thickness values of the BD-SCZ sample (n=98) and the resulting residuals were normalized using z-score transformation. We then estimated the individual joint co-variation networks of each cortical region for each patient of the BD-SCZ sample. Specifically for each individual, the intra-individual joint cortical variation between the i-th (for i = 1 to 68) and j-th (for j = 1 to 68) cortical region is equal to the inverse squared difference between the residuals of the i-th cortical region and the residuals of the j-th cortical region.

The derived individual (n=98) regional co-variation networks (n=68) were then averaged to calculate mean co-variation network maps for each cortical region (n=68) across the SCZ and BD spectrum. Next, we correlated the residuals of each cortical region (n=68) with amotivation and generated a whole brain effect size map of amotivation across the SCZ and BD spectrum. To test our hypothesis that OFC co-variation networks show spatial similarity with the regional effect size map of amotivation, we correlated each of the 68 co-variation networks with the amotivation-related cortical alteration pattern. We assessed statistical significance correcting for spatial autocorrelation using spin test^68–70^ (1000 repetitions). The specificity of this association was examined by repeating the analysis using the regional effect size map of diminished expression. To further assess the robustness of these findings repeated the analysis using independent meta-analytic cortical thickness maps derived from 4474 schizophrenia individuals from the ENIGMA consortium^40^. We generated OFC co-variation networks based on the mean cortical thickness values of the ENIGMA schizophrenia sample and compared these OFC covariation networks to the meta-analytic cortical profile associated with negative symptom severity.^40^ Please note that these meta-analytic data only include the cortical alteration patterns of total negative symptoms and did not allow to further differentiate into the two negative symptom dimensions.^40^

### Epicenter mapping of cortical alteration of amotivation

Next, we tested whether the region-specific cortico-cortical connectivity profile of the OFC or other cortical regions spatially correlates with the whole-brain cortical alteration pattern associated with amotivation severity. High spatial similarity of a region’s connectivity profile with the amotivation-related cortical alteration pattern would be indicative that this region is an putative epicenter from alterations spread to connected regions.^41,45–47,50^ We applied the epicenter mapping approach implemented in the ENIGMA Toolbox^65^ that uses normative functional and structural connectome data derived from resting-state fMRI, and diffusion MRI of unrelated healthy adults (n = 207, 83 males, mean age ± SD = 28.73 ± 3.73 years, range = 22 to 36 years) from the Human Connectome Project (HCP) (Fig.S1-2).^71,72^ Details on preprocessing and connectivity matrix generation can be found elsewhere.^70^ In brief, we spatially correlated every region’s healthy functional and structural cortico-cortical connectivity profile (derived from HCP) with the whole-brain pattern of amotivation-related cortical alteration in BD and SCZ. We repeated this approach systematically for each parcellated region with functional and structural cortical connectivity separately. Statistical significance of spatial similarity between an individual brain region’s functional (or structural) connectivity profile and amotivation-related cortical alterations was determined through spatial correlation, correcting for spatial autocorrelation with using 1000 randomly rotated cortical maps based on the spin test.^68,70^ Regions were ranked in descending order based on the strength of their correlation coefficients, with the highest-ranked regions being considered the most significant disease epicenters. A region can be considered an epicenter regardless of its own amotivation-related cortical alteration effect size, as long as it is strongly connected to other regions with high cortical alteration and weakly connected to regions with low cortical alteration.

## Results

### Demographic and clinical data

All demographic and clinical data are presented in Table 1 and in the supplementary results.

**Table 1.** Demographic and clinical data

SCZ, schizophrenia; BD, bipolar disorder I; SANS, Scale for the assessment of negative symptoms.
|  | **SCZ (N=49)** | **BD (N= 49)** | **Test-statistics** |
| --- | --- | --- | --- |
| **Age** | 36.53 +- 8.956 | 35.29 +- 9.028 | F= 0.47; P= 0.495 |
| **Sex (female/male)** | 12/37 | 21/28 | chi<sup>2</sup>= 3.701; P= 0.054 |
| **Race** |  |  |  |
| **American Indian or Alaskan Native** | 11 (22.4%) | 4 (8.2%) |  |
| **Asian** | 1 (2%) |  |  |
| **Black/African American** | 2 (4.1%) | 1 (2%) |  |
| **White** | 32 (65.3%) | 37 (75.5%) |  |
| **More than one race** | 1 (2%) | 7 (14.3%) |  |
| **SANS** |  |  |  |
| **Apathy-Avolition** | 2.69 +- 1.517 | 1.9 +- 1.388 | F= 7.343; P= 0.008 |
| **Anhedonia-Asociality** | 2.24 +- 1.493 | 1.69 +- 1.257 | F= 3.946; P= 0.05 |
| **Alogia** | 0.94 +- 1.126 | 0.16 +- 0.426 | F= 20.35; P <0.001 |
| **Blunted affect** | 1.29 +- 1.307 | 0.57 +- 1.061 | F= 8.824; P= 0.004 |
| **Amotivation** | 4.94 +- 2.764 | 3.54 +- 2.297 | F= 7.314; P= 0.008 |
| **Diminished expression** | 2.22 +- 2.266 | 0.73 +- 1.366 | F=15.533; P= <0.001 |
| **SAPS** |  |  |  |
| **Positive symptoms** | 4.92 +- 2.768 | 0.84 +- 1.419 | F= 84.381; P < 0.001 |
| **Disorganization** | 2.59 +- 2.3 | 1.6 +- 1.765 | F= 5.634; P= 0.02 |
| **HAMD-21** | 12.08 +- 9.02 | 13.8 +- 9.764 | F= 0.815; P= 0.369 |
| **Risperidone equivalent dose (mg/d)** | 7.58 +- 8.855 | 4.9 +- 10.596 | F= 1.843; P= 0.178 |

### OFC alterations and amotivation across the bipolar-schizophrenia spectrum

Spearman correlations revealed significant negative correlations of amotivation with bilateral medial OFC thickness (left: *rs* = -0.289, *p*_FDR_ = 0.008; right: *rs* = -0.221, *p*_FDR_ = 0.039) and right lateral OFC thickness (*rs* = -0.346, *p*_FDR_ = 0.002) in both disorders combined (Fig. 1). Reduced thickness of the left lateral OFC was not associated with higher amotivation across SCZ and BD (rs = -0.120, *p*_FDR_ = 0.24). Bootstrapping with 1000 resampled distributions further confirmed these associations with all 95% confidence intervals for the correlation coefficients not overlapping with zero (left medial [-0.47, -0.07]; right medial [-0.41, -0.01], right lateral [-0.53, -0.15]. In contrast to the amotivation dimension, diminished expression was not significantly correlated with reduced OFC thickness but showed a trend negative association with reduced left lateral OFC thickness (Table 2).

**Figure 1.**
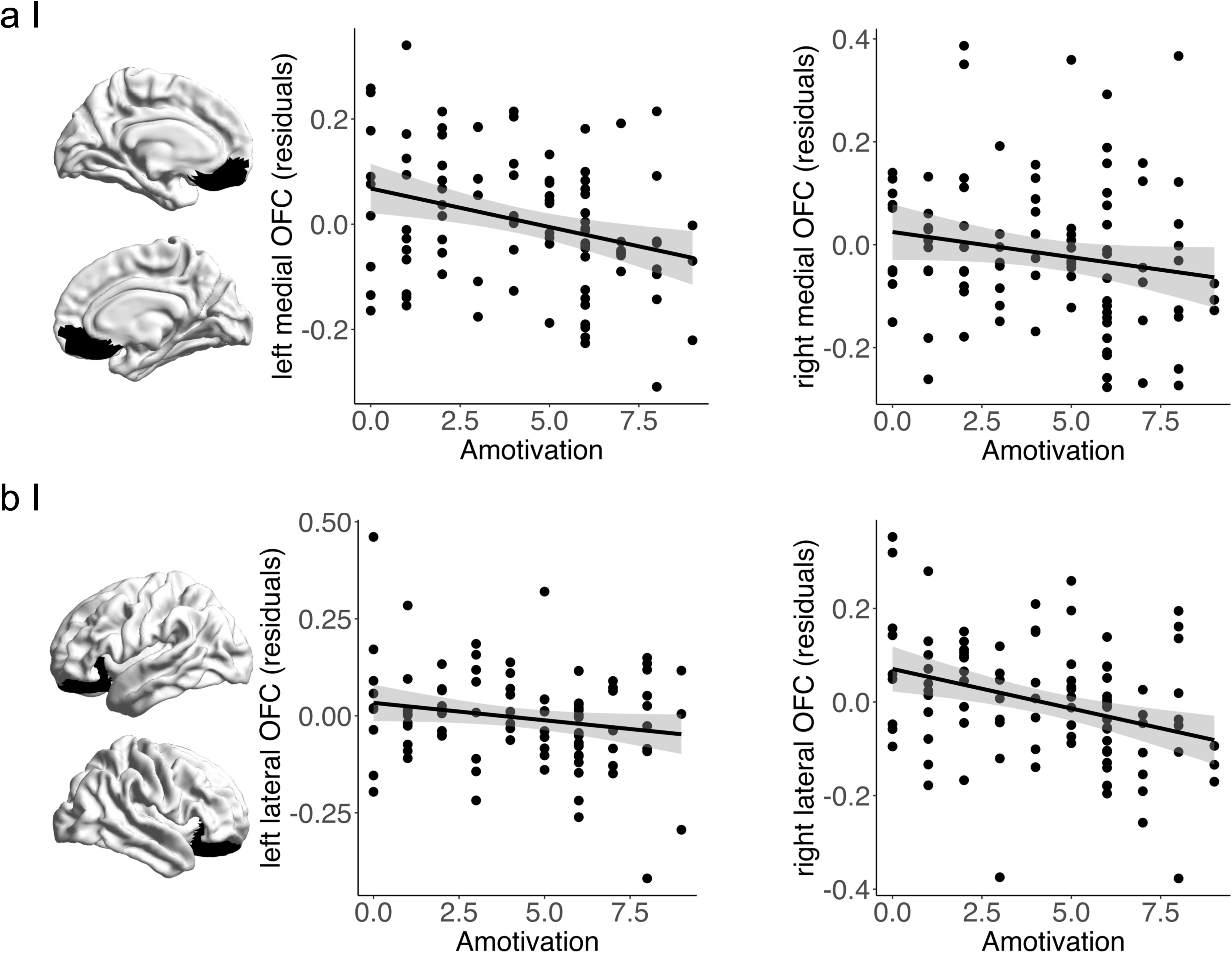
Transdiagnostic correlation analyses. **(a)** Associations between medial OFC thickness residuals (age and sex corrected) and amotivation severity. **(b)** Associations between lateral OFC thickness residuals (age and sex corrected) and amotivation severity. Error shadings correspond to standard errors. Significant associations are observed in bilateral medial OFC (left: *rs* = -0.289, *p*_FDR_ = 0.008; right: *rs* = -0.221, *p*_FDR_ = 0.039) and right lateral OFC (*rs* = -0.346, *p*_FDR_ = 0.002). In contrast, correlation analysis between amotivation and left LOFC residuals (age and sex corrected) was not significant (*rs* = -0.121, *p*_FDR_ = 0.24).

**Table 2.** Correlations of negative symptoms and OFC thickness across the BD-SCZ spectrum

FDR adjusted p value, false discovery rate adjusted p value; DimEx, diminished expression; LOFC, lateral orbitofrontal cortex; MOFC, medial orbitofrontal cortex; L, left; R, right
| **Region** | **Amotivation** | **FDR adjusted p-value** | **DimEx** | **FDR adjusted p-value** |
| --- | --- | --- | --- | --- |
| **L LOFC** | -0.121 | 0.240 | -0.194 | 0.223 |
| **R LOFC** | -0.346 | 0.002 | -0.135 | 0.264 |
| **L MOFC** | -0.289 | 0.008 | -0.131 | 0.264 |
| **R MOFC** | -0.221 | 0.039 | -0.090 | 0.380 |

To test whether associations with reduced OFC were specific for amotivation, we compared the correlation coefficients of amotivation and diminished expression with OFC thickness. We found that the correlation between right lateral OFC and amotivation was significantly different compared to the correlation between right lateral OFC and diminished expression (*Z* = -2.047, *p* = 0.021) and but not for bilateral medial OFC (left: *Z* = -1.515, *p* = 0.065; right: *Z* = -1.249, *p* = 0.108). To test whether the transdiagnostic associations were comparable in both diagnostic groups, we repeated the correlation analyses in BD and SCZ separately. The associations between higher amotivation and reduced right lateral OFC as well as bilateral medial OFC thickness were confirmed in both groups separately (Table S3). Altogether, we found that amotivation but not diminished expression was significantly associated with reduced right lateral OFC, and bilateral medial OFC thickness across BD and SCZ.

### OFC alterations and other clinical factors

We further examined whether the association of the OFC was specific for amotivation or also related to other clinical factors including risperidone equivalent dose, positive, disorganized, and depressive symptoms. Spearman correlations revealed significant negative correlations between risperidone equivalent dose and the reduced left lateral OFC (*rs* = -0.280, *p* = 0.005) as well as the right lateral OFC thickness (*rs* = -0.2, *p* = 0.046). To determine whether the observed association of amotivation and right lateral OFC thickness were confounded by antipsychotic medication, we performed partial correlation analysis including risperidone equivalent dose as covariate. The correlation between amotivation and right lateral OFC thickness remained significant after including medication dose as covariate (*rs* = -0.32, *p* = 0.002). None of the other clinical factors were significantly correlated with OFC thickness (see Table S4). In summary, medication dose showed significant correlations with bilateral lateral but not medial OFC thickness; nevertheless, correlations between amotivation and right lateral OFC thickness remained significant after controlling for medication dose.

### OFC co-variation networks and negative symptom dimensions

In a next step, we tested how OFC co-variation networks are associated with the whole-brain cortical alteration patterns (correlation coefficient) related to amotivation and diminished expression across the BD-SCZ spectrum. The derived medial OFC covariation networks reflect positive covariations with proximal regions, including the ACC, insula, and other frontal and temporal areas, as well as some distant regions, including the cuneus and lingual gyrus. Negative covariation was observed primarily in sensorimotor and parietal regions, but also in some limbic areas that are more distant from the medial OFC (Fig. 2A). For visualization of the lateral OFC covariation networks see Fig. S3.

**Figure 2.**
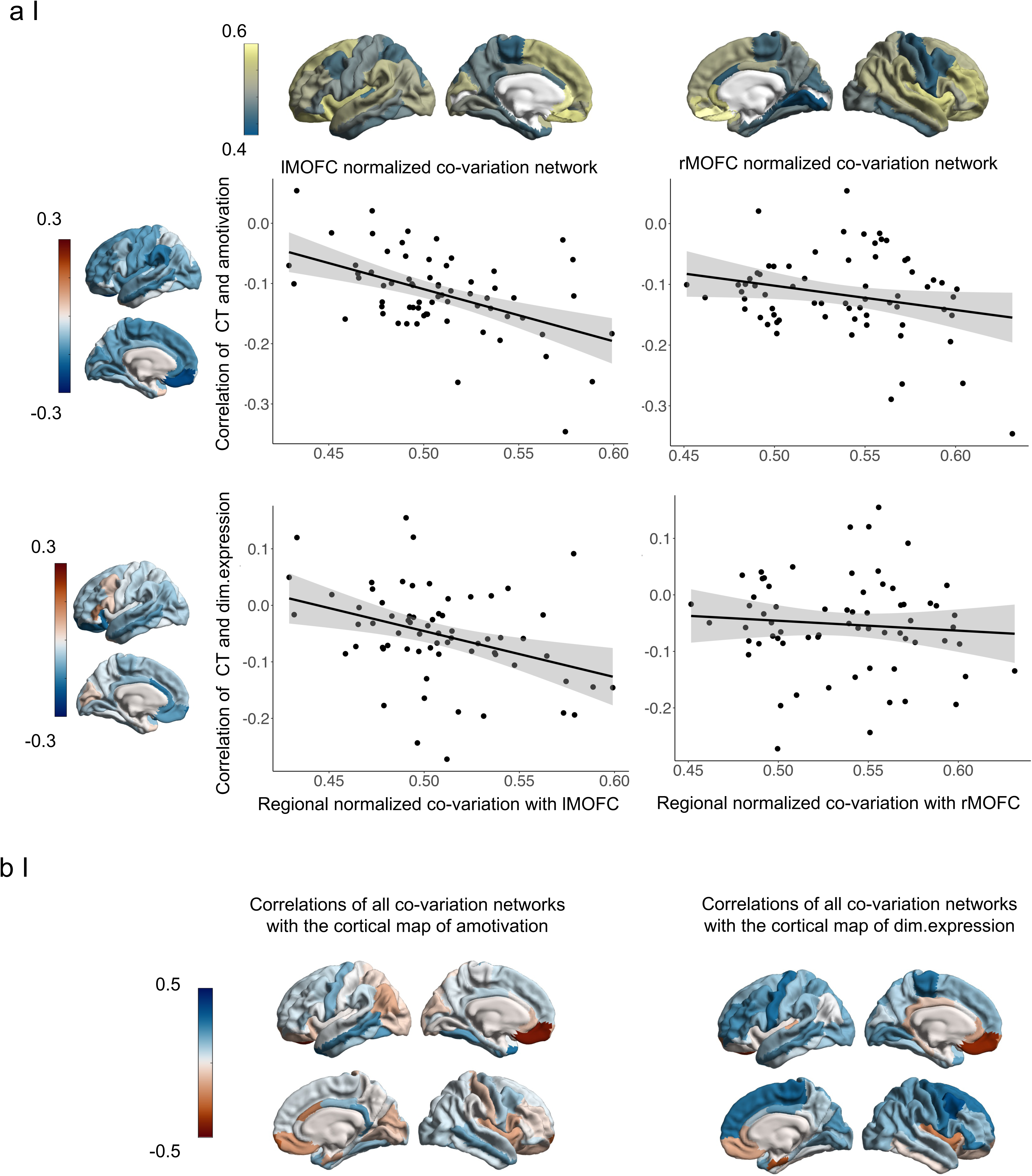
Association between co-variation networks and cortical effect sizes (correlation r) of amotivation across the BD-SCZ spectrum. **(a)** Correlations between left and right medial OFC co-variations networks and cortical effect size maps of amotivation and diminished expression respectively. Cortical regions with higher degrees of joint variation with the left medial OFC showed higher correlations with amotivation severity (*rs* = - 0.43, p_spin_ = 0.001) and diminished expression severity (*rs* = -0.38, p_spin_ = 0.004). Associations between the right medial OFC co-variation network and the cortical maps of amotivation (*rs* = -0.16,p_spin_ = 0.09) and diminished expression *rs* = -0.13, p_spin_ = 0.195) pointed in the same direction but were not significant. **(b)** Brain-wide correlations between each regional co-variation networks with the cortical alteration patterns related to amotivation and diminished expression respectively. Brain-wide comparison revealed that the associations were strongest for the left medial co-variation network.

We found that the left medial OFC co-variation network was significantly associated with the brain-wide cortical alteration pattern (correlation strength) of amotivation (*rs* = -0.43, p_spin_ = 0.001). The association with the right medial OFC co-variation network pointed in the same direction but was not significant (*rs* = -0.16, p_spin_ = 0.09) (Fig. 2A). Bilateral lateral OFC-covariation networks were not associated with the spatial pattern of cortical alterations related to amotivation severity (left/right *rs* = -0.08/-0.08, p_spin_ = 0.29/0.27) (Fig. S3). To assess whether associations with OFC co-variation networks were symptom-specific for amotivation, we repeated the analysis with the diminished expression dimension and observed comparable relationships. The cortical alteration pattern of diminished expression (correlation strength) was significantly associated with the spatial pattern of the left medial OFC co-variation network (*rs* = 0.38, p_spin_ =0.004) but not with the right medial OFC co-variations network (*rs* = -0.13, p_spin_ = 0.19) (Fig. 2A). Bilateral lateral OFC networks were not associated with the spatial pattern of cortical alterations related to diminished expression severity (left/right *rs* = -0.003/-0.06, p_spin_ = 0.47/0.31, Fig. S3). We next assessed the correlation strengths of the left medial OFC co-variation network compared to other regional co-variation networks and systematically repeated the spatial correlation of the cortical alterations of amotivation with all other cortical co-variation networks (n= 68 co-variation networks) (Fig. 2C). Ranking the correlation strengths, we found that the left medial OFC co-variation network ranked first (Fig. 2C, Table S8). Brain-wide comparison of the association between left medial OFC co-variation network and the cortical alteration pattern of diminished expression mirrored these findings (Fig. 2C, Table S9). Lastly, to show robustness of the association between OFC co-variation networks and the spatial cortical thickness pattern of amotivation and diminished expression, we repeated the analysis using independent OFC co-variation networks and independent cortical alteration patterns related to total negative symptoms (including both negative symptom dimensions). To this end, we repeated the analysis using meta-analytic data from 4474 individuals with schizophrenia from the ENIGMA consortium.^40^ In the ENIGMA schizophrenia sample (n=4474 correlating the OFC co-variation networks with the spatial pattern related to total negative symptom severity revealed significant associations with bilateral lateral OFC co-variation networks (left/right *rs* = -0.27/-0.25, p_spin_ = 0.04/0.05). No significant associations were observed for the medial OFC co-variation networks (left/right *rs* = -0.08/0.05, p_spin_ = 0.31/0.37). Taken together, we observed that OFC co-variation networks were spatially associated with the cortical alteration pattern related to negative symptom severity, unspecific to either the amotivation or diminished expression dimension. In a transdiagnostic BD-SCZ spectrum sample this association specifically emerged for the left medial OFC co-variation network. In an independent large-scale schizophrenia sample from the ENIGMA consortium the association was observed in the left and right lateral OFC co-variation networks.

### Epicenter mapping of cortical alteration associated with negative symptom dimensions

We finally examined how the cortical alteration pattern associated with amotivation was spatially correlated to the normative functional and structural connectivity of the OFC and other cortical regions. With regards to cortico-cortical connectivity of the OFC, we found that structural connectivity of the right medial OFC (r_struc_=0-.275, p_spin_=0.018) and left lateral OFC (r_struc_=0-.251, p_spin_=0.003) were significantly correlated with the whole-brain cortical alteration profile of amotivation severity (Fig. 3A, Tables S10-11). Thus, cortical regions with strong connections to the right medial OFC and left lateral OFC showed relatively higher cortical thickness reduction in relation to amotivation severity. Beyond OFC connectivity, the right rostral anterior cingulate cortex (rACC) emerged as the strongest epicenter of the amotivation-related cortical alteration pattern irrespectively of using functional or structural cortico-cortical connectivity (r_fun_=-0.295, p_spin_=0.0098, r_struc_=0-.410, p_spin_=0.001, Fig. 3A, Tables S10-11). Additionally significant functional and structural epicenters included other prefrontal and limbic areas adjacent to the OFC including bilateral frontal poles, the right caudal ACC, right PCC, and the right superior frontal gyrus (Fig.3A, Table S10-11). Exploring the associations between region-specific cortical-connectivity and the spatial cortical pattern related to diminished expression, we identified only the right temporal pole (r_struc_=0-.348, p_spin_=0.005) and the left medial OFC (r_struc_=0-.217, p_spin_=0.045) as putative epicenters (Fig. 3B, Table S12-13). Taken together, the right medial and left lateral OFC as well as adjacent regions in particular the right rACC emerged as the most important epicenter of the amotivation-related cortical alteration profile. These epicenters were not significantly associated with the cortical alteration pattern related to diminished expression. Overall, the cortical alteration pattern associated with amotivation was spatially more strongly anchored to region-specific cortico-cortical connectivity than the cortical alteration pattern associated with diminished expression.

**Figure 3.**
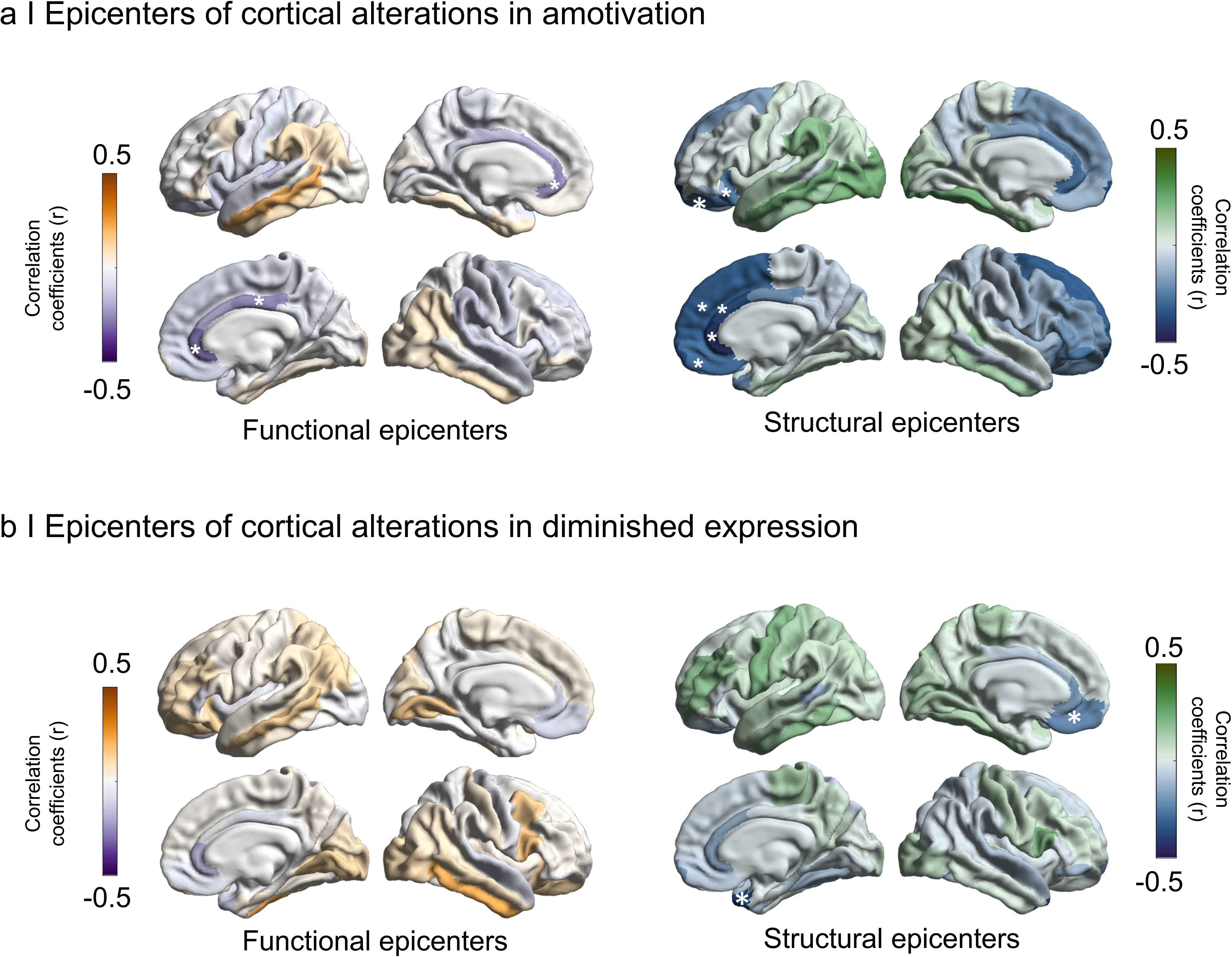
Epicenters of cortical alteration patterns of amotivation and diminished expression. Spatial correlations between cortical alteration patterns associated with **(a)** amotivation and **(b)** diminished expression and seed-based cortico-cortical connectivity of each region with all other regions were used to identify epicenters of symptom-specific cortical alterations. Epicenters are regions whose connectivity profiles significantly spatially correlated with the cortical alteration map related to each negative symptom dimension. We assessed statistical significance using spin permutation tests (1000 repetitions) and repeated this procedure systematically to assess the epicenter value of every cortical region using both functional and structural connectivity matrices. **(a)** Correlation coefficients indexing spatial similarity between the amotivation-related cortical alteration pattern and seed-based functional (left) and structural (right) connectivity measures for every cortical region. The five most significant epicenters are highlighted with asterisks. Functional epicenters included bilateral rACC, right PCC. Structural epicenters included bilateral frontal pole cortex, right rACC, cACC, mOFC, and superiorfrontal cortex. **(b)** Correlation coefficients indexing spatial similarity between the diminished expression-related cortical alteration pattern and seed-based functional (left) and structural (right) connectivity measures for every cortical region. Significant epicenters are highlighted with asterisks. No significant functional epicenters were identified. Structural epicenters included the right temporal pole cortex, and the left mOFC.

## Discussion

Leveraging a transdiagnostic approach, we tested the hypotheses that OFC thickness alteration and OFC co-variation networks are related to amotivation severity across the BD-SCZ spectrum. We found that reduced right lateral OFC and bilateral medial OFC thickness were specifically associated with amotivation but not with diminished expression. This relationship was not driven by other clinical factors and was also replicated within the BD and SCZ cohort separately. We further observed negative associations between the left medial OFC co-variation network and cortex-wide alteration patterns related to amotivation and diminished expression severity. These findings suggest that OFC co-variation might be implicated in both negative symptom dimensions. Confirmatory analyses in independent meta-analytic data from the ENIGMA consortium revealed similar associations between total negative symptom severity and lateral OFC co-variation networks. This indicates that the relationship between negative symptoms and cortical thickness reduction is not randomly distributed across the cortex but anchored to structural co-variation profiles of the OFC. Finally, we found that the cortico-cortical connectivity profiles of the right medial and left lateral OFC as well as adjacent regions including the right rACC were spatially associated with the cortical alteration related to amotivation severity. Collectively, this study identified OFC alterations as transdiagnostic morphometric signature of amotivation, and provide insights into network mechanisms underlying the brain-wide cortical alteration pattern of negative symptoms across SCZ and BD.

Given the OFC’s pivotal role in decision-making processes,^73^ such as expected value and effort-cost computation, alterations in this area are widely recognized as important factor for motivational deficits in SCZ.^19,20,74^ Gold and colleagues initially demonstrated that impaired value representation is related to higher NS^18^ and more recently found that these impairments extend to maintaining reward-related information over time.^75^ Extending this work, several studies have shown that impaired value and effort-cost computation are associated with more severe negative symptoms^76^ and particular motivational deficits.^77–81^ Motivational deficits have been further linked to blunted OFC fMRI signals during reinforcement learning in SCZ^27^ and intertemporal decision-making across SCZ but also BD and major depressive disorders.^82^ Our data extend these behavioral and fMRI findings by providing a transdiagnostic link between neuroanatomical alterations of the OFC and motivational impairments across BD and SCZ. The observed relationships between lateral and medial OFC thickness reduction and amotivation scores is consistent with previous reports examining total negative symptoms in individuals with chronic SCZ^37^ and first-episode psychosis.^34^ By deconstructing different negative symptoms, our findings suggest a stronger association between OFC thickness reduction and the amotivation compared to the diminished expression dimension. It should be noted however, that previous work has reported comparable associations of OFC alterations with both negative symptom dimensions in first-episode psychosis.^34^ Rather than being mutually exclusive, these results may provide a complementary picture of the involvement of the OFC in the pathogenesis of amotivation and diminished expression. The OFC is embedded in multiple cortico-cortical and cortico-subcortical networks and participates in a plethora of complex functions, including the processing of emotions.^73,83–85^ Alterations in the OFC may therefore represent both a key substrate of motivational deficits within the BD-SCZ spectrum, as well as play a role in the development of diminished emotional expressivity. However, contrary to the accumulating evidence supporting compromised OFC contributions to decision-making and reward learning in association with motivational deficits, studies linking impaired OFC signaling during emotional processing within the SCZ spectrum are relatively scarce.^19,20,74^ Thus, future research should combine decision-making and emotion-processing tasks with multimodal neuroimaging to disentangle how distinct OFC abnormalities contribute to both motivational deficits and diminished expressivity across the psychosis continuum.

We further identified that the brain-wide cortical alterations associated with amotivation severity were not randomly distributed but constrained within the co-variation network of the left medial OFC. This association was not specific for amotivation but also observed with the cortical thickness reduction pattern related to diminished expression suggesting a general mechanism for both negative symptom dimensions. In line with these findings confirmatory analysis using independent meta-analytic data revealed similar associations between the lateral OFC co-variation networks and the cortical alteration pattern associated with total negative symptom severity. The discrepancy between the transdiagnostic findings in our BD-SCZ sample and the meta-analytic data derived from a recent ENIGMA publication might be due to the differences in study samples and assessments of negative symptoms. Cortical covariance networks describe coordinated changes in cortical thickness across different brain regions,^66^ and may reflect maturational coupling and synchronized trophic processes during cortical development.^86,87^ They appear to be organized across multiple scales, such as genetic correlations between regions and hierarchical axis of cortical organization.^64^ Moreover, it has been demonstrated that these networks describe coordinated patterns of cortical atrophy in different neuropsychiatric disorders, including schizophrenia.^42,66,88^ Recent studies have further shown that circumscribed covariance networks are associated with distinct symptom dimensions.^43,44^ Expanding on these insights, our results suggest that circumscribed cortical co-variation networks may represent the patterns of cortical alterations associated with distinct symptom profiles. More specifically, we observed that medial OFC co-variation networks align with cortical patterns related to amotivation and diminished expressivity within the BD-SCZ spectrum. In addition, our analysis using meta-analytic data points toward a role of lateral OFC co-variation networks for total negative symptoms in SCZ. One possible explanation for the observed associations might be that genetic factors and trophic processes that guide medial and lateral OFC co-variation networks are also implicated in the pathophysiology of cortical alterations associated with negative symptoms. Therefore, understanding the cellular and molecular mechanisms driving the evolution of large-scale structural co-variance networks might be key to identify disrupted biological systems underlying negative symptoms in the BD-SCZ spectrum.^66^ We finally contextualized the contribution of normative brain network architecture to the spatial pattern of cortical alterations associated with amotivation in BD and SCZ. A growing body of literature provides evidence that cross-disorder and disease-specific cortical alteration patterns of neuropsychiatric disorders are guided by normative connectivity profiles of distinct cortical regions.^89^ In schizophrenia, temporo-paralimbic regions have been identified as putative epicenters whose connectivity profile spatially constrains the extent of cortical atrophy.^45,46,89^ Analysis across different disease stages further suggest that epicenters of cortical atrophy emerged first in occipital regions early in the illness and shifts to prefrontal regions with illness progression.^45,89^ Cross-disorder comparison of BD and SCZ revealed convergence of parieto-temporal and frontal epicenters of cortical alterations seen in both disorders. Furthermore, higher psychotic symptom severity has been shown to be associated with sensory-motor and paralimbic epicenters suggesting that individual clinical facets contribute to the development of distinct symptom-specific epicenters.^45^ Extending these observations, our findings indicate that the cortical alteration pattern associated with amotivation severity in BD and SCZ is anchored to the connectivity profiles of prefrontal and limbic areas. The right medial OFC, left lateral OFC and particular right rACC emerged as significant epicenters of the cortical alteration pattern associated with amotivation. The rACC is tightly connected to proximal regions, in particular the OFC, and other areas of the ACC and prefrontal cortex.^90^ Additionally, it is also a major hub of large-scale networks such as the default mode network.^91^ This position at the intersection between motivation and cognitive control networks aligns with the important function of the rACC in value-based decision making.^92–94^ Altered structure and function of the ACC has been consistently reported to be associated with motivational deficits across neuropsychiatric disorders including schizophrenia and affective disorders.^20,21,31^ Our findings demonstrate that the spatial pattern of cortical alterations associated with motivational deficits is not randomly distributed but is determined by the functional and structural connectivity profile of the OFC and ACC. This is consistent with the growing recognition that pathological processes in psychiatric disorders follow network principles and are related to the underlying connectome architecture.^41,95,96^ Although it is not possible to deduce such mechanisms from the present findings, one hypothesis could be that the OFC and ACC act as gateways from which altered neurodevelopment or neurodegeneration propagates to the connected areas.

### Limitations and future directions

Capitalizing on BD-SCZ spectrum data, we provide support for transdiagnostic orbitofrontal morphometric signatures of motivational negative symptoms and network characteristics linked to the cortical pattern associated with negative symptoms. However, larger BD-SZ spectrum samples including different stages of both conditions are needed (i) to replicate our findings and (ii) to study the role of localized and network alterations of the OFC and ACC during early disease courses. Although a harmonized assessment of negative symptoms was used by applying the SANS to both diagnostic groups a multimodal assessment including newer scales, self-reports and behavioral measures would be valuable for future research. The severity of the amotivation dimension and diminished expression were comparable to previous psychometric and neuroimaging studies in the BD-SCZ spectrum,^13–16,34^ therefore allowing comparison with the existing literature. However, defining a priori thresholds of negative symptom severity might help to enable examining brain-behavior relationships across the full range of negative symptom severity.

## Conclusion

Leveraging a combined BD-SCZ sample, we found that reduced OFC thickness reflects a transdiagnostic signature of amotivation severity. Notably, these associations were not related to other symptoms or specific to one diagnostic group, highlighting its potential utility as target for transdiagnostic patient stratification and treatment development. Our results further suggest a region-specific link between medial OFC co-variation networks and cortical patterns of amotivation and diminished expression. More broadly, our work contributes to the growing recognition that neuropsychiatric psychopathologies can be characterized by alterations in circumscribed cortical networks that are linked to distinct network features of the brain’s connectome architecture.

## Data Availability

All data produced in the present study are available upon reasonable request to the authors

## Acknowledgements

All data used in this study were derived from the UCLA CNP cohort. Data can be download from the publicly available database OpenfMRI (https://openfmri.org/dataset/ds000030/). BTTY is supported by the NUS Yong Loo Lin School of Medicine (NUHSRO/2020/124/TMR/LOA), the Singapore National Medical Research Council (NMRC) LCG (OFLCG19May-0035), NMRC CTG-IIT (CTGIIT23jan-0001), NMRC STaR (STaR20nov-0003), Singapore Ministry of Health (MOH) Centre Grant (CG21APR1009), the Temasek Foundation (TF2223-IMH-01), and the United States National Institutes of Health (R01MH120080 & R01MH133334). EW receives funding from UK Research and Innovation (UKRI) under the UK government’s Horizon Europe / ERC Frontier Research Guarantee [BrainHealth, grant number EP/Y015037/1]. Research reported in this publication was supported by the National Institutes of Mental Health under Award Number R01MH121246.

