## Supplement for "Orbitofrontal thickness and network associations as transdiagnostic signature of negative symptoms along the bipolar-schizophrenia spectrum"

**Altered OFC structure and interregional covariance as transdiagnostic signature of amotivation along the bipolar-schizophrenia spectrum**

**Supplement**

**Methods**

**Image Acquisition and Processing**

Imaging data were collected with one of two 3T Siemens Trio scanners. One was located at the Ahmanson-Lovelace Brain Mapping Center (Siemens version syngo MR B15) and the other one at the Staglin Center for Cognitive Neuroscience (Siemens version syngo MR B17) at UCLA. High-resolution anatomical scans were collected with a T1-weighted matched-bandwidth sequence with the following parameters: 4 mm slices, TR/TE=5000/34 ms, 4 averages, matrix=128×128. 90 degree flip angle. We used FreeSurfer Version 5.3.0 recon-all function to perform cortical reconstruction and volumetric segmentation. The automated recon-all function processing streamline for structural MRI data includes motion correction, removal of non-brain tissue, automated Talairach transformation, segmentation of the subcortical white matter and deep gray matter volumetric structures, intensity normalization, tessellation of the gray matter-white matter boundary, topology correction, and surface generation and deformation. Quality Control (QC) was performed using standard ENIGMA QC protocols (http://enigma.ini.usc.edu/protocols/imaging-protocols).

**Group Comparison**

Group comparisons of standardized residuals of OFC thickness from HC, BD, SCZ (total *n*= 220) were performed using one-way univariate analysis of covariance (ANCOVA). T-tests were used for subsequent post hoc analyses (IBM SPSS Version 28.0.1.1 (14). Results can be found in Table S2.

**Results**

**Demographic and clinical data**

All demographic and clinical data are presented in Table 1. There was no group difference in age [*F(1, 96)* = 0.47, *p* = 0.495], but trend-level higher proportion of male subjects in the SCZ (75%) group compared to the BD group (57%, chi2 =3.701, p =0.054). In both SCZ and BD, levels of negative symptoms were mild to moderate, with higher amotivation [*F(1, 95)* = 7.31, *p* = 0.008] and diminished expression severity [*F(1, 96)* = 15.53, *p* = <0.001] in SCZ compared to BD. With respect to the other symptom dimensions, we found higher SAPS positive symptom dimension scores [*F(1,96)* = 84.381, *p* < 0.001] as well as higher levels in SAPS disorganization [*F(1,94)* = 5.634; *p* = 0.02] in the SCZ group than the BD group. In contrast, HAMD-21 scores did not differ between groups [*F(1,96)* = 0.815; *p* = 0.369]. Regarding antipsychotic medication dose, we found no significant difference between the SCZ and the BD group [*F(1,96)* = 1.843; *p* = 0.178].

Table S1. Mean values of cortical thickness across all datasets

| Region | HC (n= 122) | BD (n=49) | SCZ (n=49) |
| --- | --- | --- | --- |
| L LOFC | 2.49 (0.12) | 2.48 (0.14) | 2.46 (0.134) |
| R LOFC | 2.42 (0.122) | 2.4 (0.145) | 2.38 (0.14) |
| L MOFC | 2.3 (0.123) | 2.29 (0.143) | 2.30 (0.131) |
| R MOFC | 2.24 (0.112) | 2.16 (0.146) | 2.19 (0.162) |

Table S1: Data are presented as means and standard deviations of cortical thickness (mm). HC, healthy controls; BD, patients with bipolar disorder; SZ, patients with schizophrenia; LOFC, lateral orbitofrontal cortex; MOFC, medial orbitofrontal cortex; L, left; R, right

Table S2. Group differences in standardized residuals of OFC thickness

| Region | HC (n= 122) | BD (n=49) | SCZ (n=49) | df | Error | F | p-value | Post-hoc test^a^ | Post-hoc p-value^a^ |
| --- | --- | --- | --- | --- | --- | --- | --- | --- | --- |
| L LOFC | 0.0032 | 0.009 | -0.0165 | 2 | 217 | 0.656 | 0.520 |  |  |
| R LOFC | 0.001 | 0.006 | -0.009 | 2 | 217 | 0.205 | 0.815 |  |  |
| L MOFC | -0.004 | -0.004 | 0.014 | 2 | 217 | 0.399 | 0.672 |  |  |
| R MOFC | 0.014 | -0.039 | 0.003 | 2 | 217 | 3.19 | **0.043** | HC>BD  HC=SCZ  BD=SCZ | **0.038**  0.593  0.197 |

Table S2. Data are presented as standardized residuals, age and sex regressed-out for orbitofrontal cortical thickness. Standard error in parenthesis. Univariate analyses were corrected for site. Mean values of cortical thickness (mm) are presented in Table 2. HC, healthy controls; BD, patients with bipolar disorder; SZ, patients with schizophrenia; LOFC, lateral orbitofrontal cortex; MOFC, medial orbitofrontal cortex; L, left; R, right; Significant group differences in bold. ^a^Post-hoc tests are corrected for multiple comparisons using the Holm-Bonferroni method (FWE p<0.05).

Table S3. Correlation amotivation and OFC in BD and SCZ separately

| Region | BD Amotivation | p-value | SCZ Amotivation | p-value |
| --- | --- | --- | --- | --- |
| L LOFC | -0.118 | 0.42 | -0.107 | 0.464 |
| R LOFC | **-0.371** | 0.009 | **-0.323** | 0.024 |
| L MOFC | **-0.322** | 0.026 | **-0.342** | 0.016 |
| R MOFC | **-0.298** | 0.040 | **-0.239** | 0.099 |

Table S3. BD, bipolar disorder; SCZ, schizophrenia; LOFC, lateral orbitofrontal cortex;

MOFC, medial orbitofrontal cortex; L, left; R, right.

Table S4. Correlations between other clinical factors and OFC across the BD-SCZ spectrum

| Region | Risp eq dose | p-value | Positive symptoms | p-value | Disorganization | p-value | HAMD-21 | p-value |
| --- | --- | --- | --- | --- | --- | --- | --- | --- |
| L LOFC | **-0.280** | 0.005 | 0.016 | 0.687 | -0.022 | 0.755 | 0.053 | 0.365 |
| R LOFC | **-0.201** | 0.046 | 0.034 | 0.560 | 0.022 | 0.814 | -0.069 | 0.610 |
| L MOFC | -0.062 | 0.476 | 0.017 | 0.747 | -.132 | 0.242 | -0.021 | 0.799 |
| R MOFC | 0.010 | 0.959 | -0.016 | 0.820 | -0.161 | 0.144 | -0.154 | 0.223 |

Table S4. risp eq dose, risperidone equivalent dose, LOFC, lateral orbitofrontal cortex; MOFC, medial orbitofrontal cortex; L, left; R, right

Fig S1. Normative functional degree centrality derived from the HCP sample (n=207)

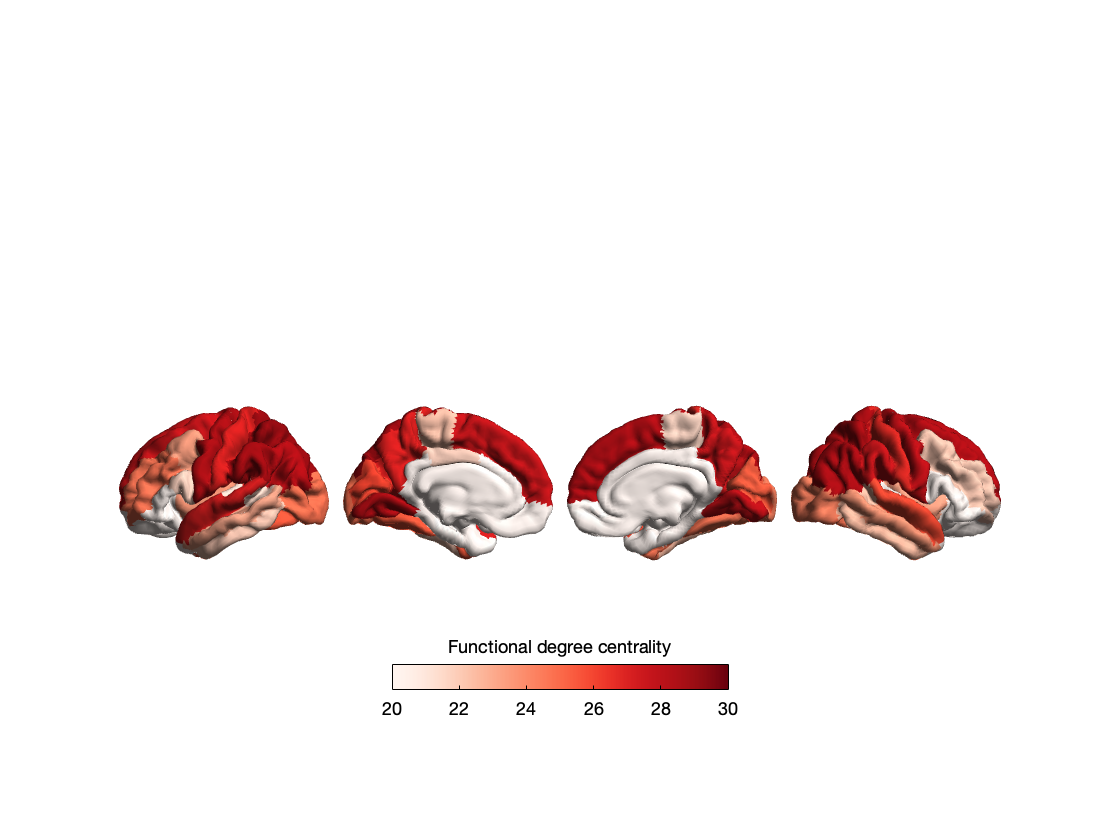

Fig S2. Normative structural degree centrality derived from the HCP sample (n=207)

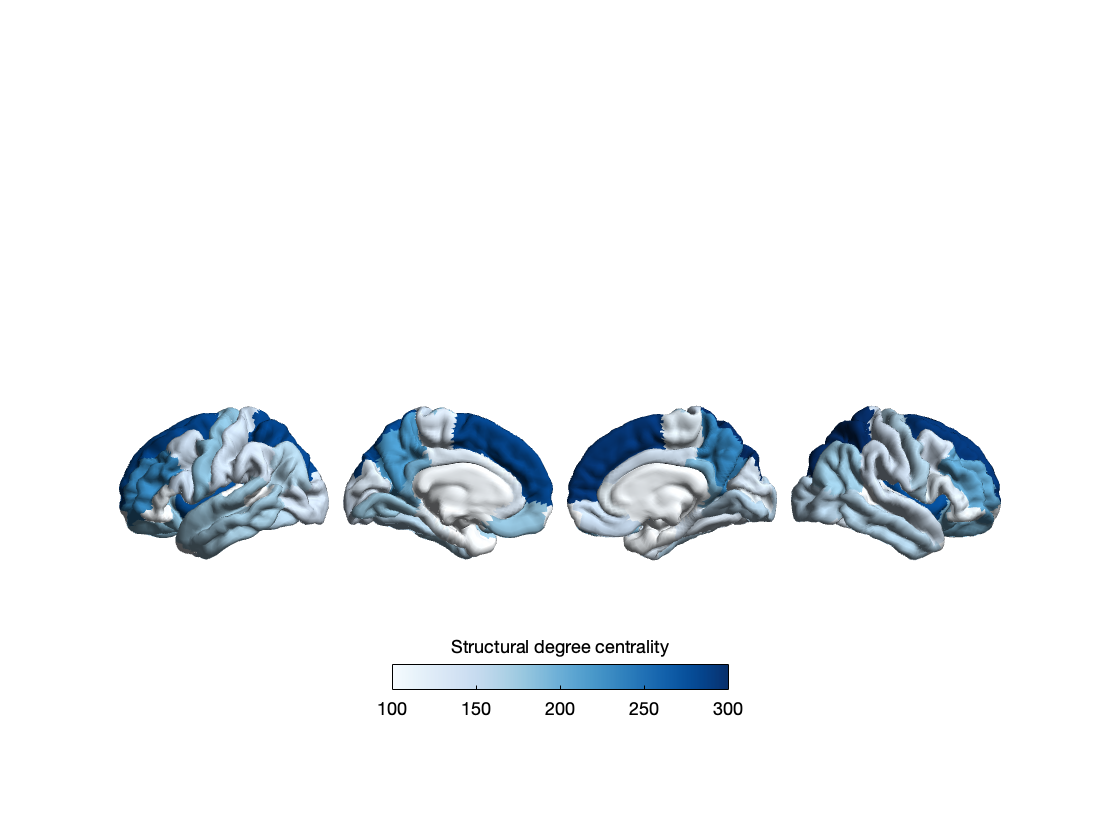

Fig. S3. Association between co-variation networks and cortical effect sizes (correlation r) of amotivation and diminished expression across the BD-SCZ spectrum

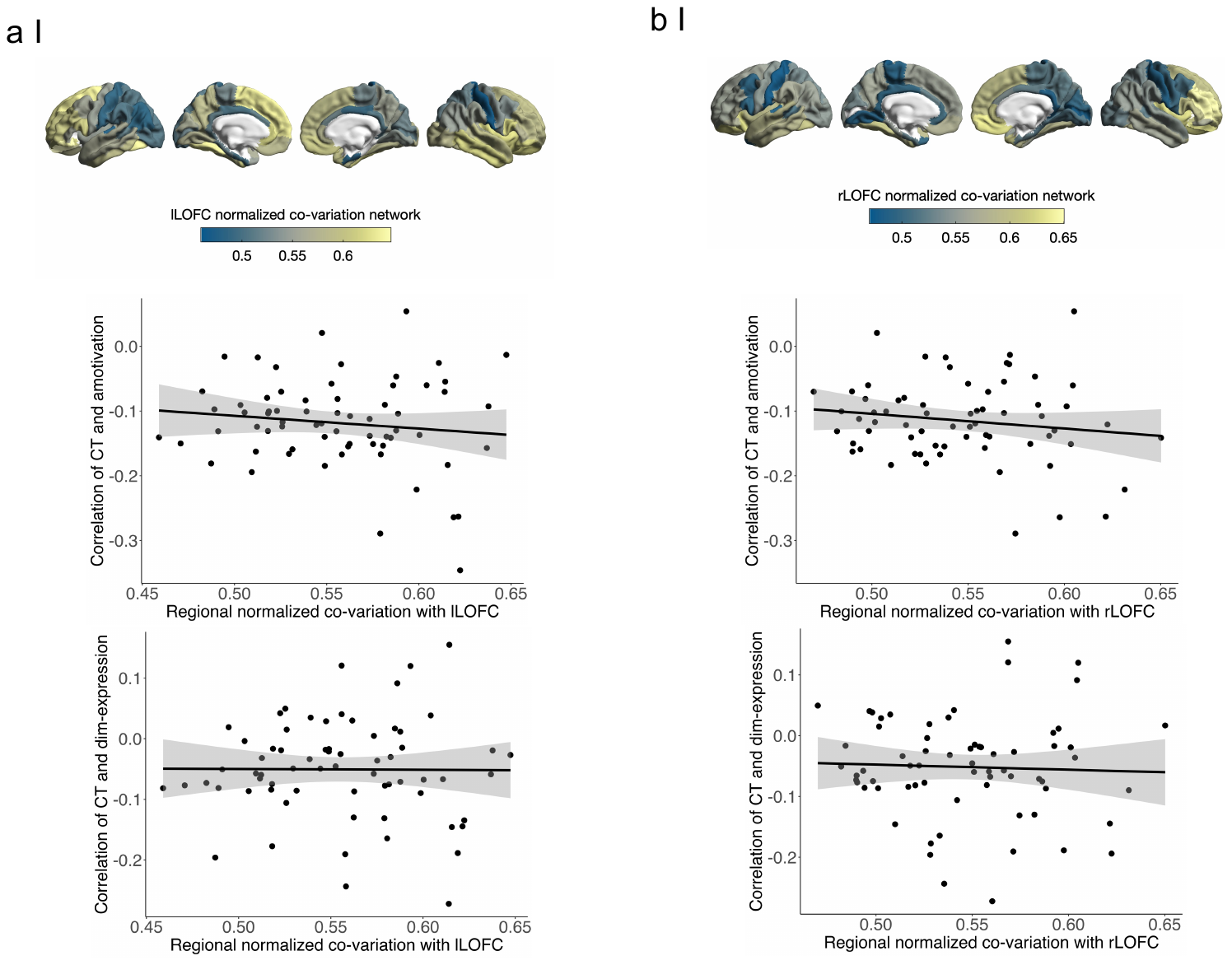

**Figure S3.** Association between co-variation networks and cortical effect sizes (correlation r) of amotivation and diminished expression across the BD-SCZ spectrum. **(a)** Correlations between the left lOFC co-variations network and the cortical effect size maps of amotivation and diminished expression respectively. The left lOFC co-variation network did not show significant spatially correlation with the cortical alteration pattern of amotivation or diminished expression (amotivation, rs = -0.080, p_spin_ = 0.29; diminished expression, rs = -0.003, p_spin_ = 0.47). **(b)** Correlations between the right lOFC co-variation network and the cortical effect size maps of amotivation and diminished expression respectively. Similar to the results with the left lOFC co-variation network, the right lOFC co-variation network did not show significant correlations with the cortical alteration pattern of amotivation or diminished expression (amotivation, rs = -0.090, p_spin_ = 0.27; diminished expression, rs = -0.06, p_spin_ = 0.31).

Table S8. Spatial correlation of co-variation networks with amotivation-related cortical alterations

| DKT_Region | r_value | pspin | Rank |
| --- | --- | --- | --- |
| L_medialorbitofrontal_thickavg | -0.425 | 0.001 | 1 |
| R_frontalpole_thickavg | -0.285 | 0.010 | 2 |
| R_caudalanteriorcingulate_thickavg | -0.202 | 0.053 | 3 |
| R_medialorbitofrontal_thickavg | -0.163 | 0.089 | 4 |
| R_insula_thickavg | -0.142 | 0.113 | 5 |
| L_inferiorparietal_thickavg | -0.127 | 0.159 | 6 |
| R_entorhinal_thickavg | -0.124 | 0.160 | 7 |
| R_parsopercularis_thickavg | -0.122 | 0.197 | 8 |
| R_cuneus_thickavg | -0.111 | 0.195 | 9 |
| L_frontalpole_thickavg | -0.100 | 0.201 | 10 |
| R_pericalcarine_thickavg | -0.086 | 0.195 | 11 |
| R_lateralorbitofrontal_thickavg | -0.085 | 0.272 | 12 |
| L_lateralorbitofrontal_thickavg | -0.078 | 0.290 | 13 |
| R_postcentral_thickavg | -0.076 | 0.257 | 14 |
| L_rostralanteriorcingulate_thickavg | -0.074 | 0.273 | 15 |
| L_cuneus_thickavg | -0.061 | 0.327 | 16 |
| R_lingual_thickavg | -0.053 | 0.279 | 17 |
| L_lateraloccipital_thickavg | -0.045 | 0.370 | 18 |
| L_superiorparietal_thickavg | -0.027 | 0.395 | 19 |
| R_rostralmiddlefrontal_thickavg | -0.024 | 0.463 | 20 |
| R_transversetemporal_thickavg | -0.022 | 0.423 | 21 |
| L_transversetemporal_thickavg | 0.001 | 0.567 | 22 |
| L_caudalanteriorcingulate_thickavg | 0.004 | 0.442 | 23 |
| L_insula_thickavg | 0.004 | 0.459 | 24 |
| R_precuneus_thickavg | 0.005 | 0.464 | 25 |
| L_lingual_thickavg | 0.005 | 0.487 | 26 |
| R_parstriangularis_thickavg | 0.010 | 0.437 | 27 |
| R_supramarginal_thickavg | 0.011 | 0.474 | 28 |
| L_postcentral_thickavg | 0.017 | 0.459 | 29 |
| L_paracentral_thickavg | 0.019 | 0.450 | 30 |
| R_rostralanteriorcingulate_thickavg | 0.020 | 0.396 | 31 |
| L_parsorbitalis_thickavg | 0.021 | 0.411 | 32 |
| L_parsopercularis_thickavg | 0.027 | 0.400 | 33 |
| L_superiortemporal_thickavg | 0.029 | 0.418 | 34 |
| L_precuneus_thickavg | 0.034 | 0.381 | 35 |
| R_superiorfrontal_thickavg | 0.036 | 0.340 | 36 |
| R_paracentral_thickavg | 0.038 | 0.343 | 37 |
| R_superiorparietal_thickavg | 0.042 | 0.405 | 38 |
| R_inferiortemporal_thickavg | 0.045 | 0.336 | 39 |
| L_pericalcarine_thickavg | 0.047 | 0.348 | 40 |
| L_isthmuscingulate_thickavg | 0.049 | 0.326 | 41 |
| R_parahippocampal_thickavg | 0.052 | 0.381 | 42 |
| R_inferiorparietal_thickavg | 0.052 | 0.339 | 43 |
| L_entorhinal_thickavg | 0.052 | 0.328 | 44 |
| L_rostralmiddlefrontal_thickavg | 0.052 | 0.352 | 45 |
| R_superiortemporal_thickavg | 0.053 | 0.350 | 46 |
| L_superiorfrontal_thickavg | 0.057 | 0.346 | 47 |
| R_parsorbitalis_thickavg | 0.057 | 0.264 | 48 |
| L_supramarginal_thickavg | 0.059 | 0.277 | 49 |
| R_lateraloccipital_thickavg | 0.068 | 0.322 | 50 |
| L_posteriorcingulate_thickavg | 0.070 | 0.243 | 51 |
| L_caudalmiddlefrontal_thickavg | 0.072 | 0.271 | 52 |
| L_parahippocampal_thickavg | 0.078 | 0.237 | 53 |
| R_precentral_thickavg | 0.088 | 0.169 | 54 |
| L_inferiortemporal_thickavg | 0.094 | 0.207 | 55 |
| L_parstriangularis_thickavg | 0.100 | 0.215 | 56 |
| R_caudalmiddlefrontal_thickavg | 0.110 | 0.168 | 57 |
| R_posteriorcingulate_thickavg | 0.121 | 0.128 | 58 |
| R_bankssts_thickavg | 0.128 | 0.128 | 59 |
| R_fusiform_thickavg | 0.128 | 0.135 | 60 |
| R_temporalpole_thickavg | 0.147 | 0.108 | 61 |
| L_precentral_thickavg | 0.151 | 0.080 | 62 |
| R_middletemporal_thickavg | 0.163 | 0.090 | 63 |
| L_bankssts_thickavg | 0.163 | 0.101 | 64 |
| R_isthmuscingulate_thickavg | 0.194 | 0.054 | 65 |
| L_fusiform_thickavg | 0.198 | 0.045 | 66 |
| L_middletemporal_thickavg | 0.216 | 0.031 | 67 |
| L_temporalpole_thickavg | 0.272 | 0.016 | 68 |

Table S9. Spatial correlation of co-variation networks with diminished expression-related cortical alterations

| DKT_Region | r_value | pspin | Rank |
| --- | --- | --- | --- |
| L_medialorbitofrontal_thickavg | -0.385 | 0.004 | 1 |
| R_entorhinal_thickavg | -0.301 | 0.017 | 2 |
| R_insula_thickavg | -0.223 | 0.042 | 3 |
| L_frontalpole_thickavg | -0.209 | 0.056 | 4 |
| L_transversetemporal_thickavg | -0.145 | 0.142 | 5 |
| R_medialorbitofrontal_thickavg | -0.131 | 0.195 | 6 |
| L_rostralanteriorcingulate_thickavg | -0.120 | 0.158 | 7 |
| R_frontalpole_thickavg | -0.108 | 0.212 | 8 |
| L_isthmuscingulate_thickavg | -0.097 | 0.195 | 9 |
| R_rostralanteriorcingulate_thickavg | -0.097 | 0.240 | 10 |
| R_lateralorbitofrontal_thickavg | -0.059 | 0.311 | 11 |
| L_posteriorcingulate_thickavg | -0.058 | 0.299 | 12 |
| L_parahippocampal_thickavg | -0.049 | 0.330 | 13 |
| R_transversetemporal_thickavg | -0.041 | 0.370 | 14 |
| R_inferiortemporal_thickavg | -0.021 | 0.443 | 15 |
| L_lateralorbitofrontal_thickavg | -0.003 | 0.469 | 16 |
| L_insula_thickavg | 0.001 | 0.462 | 17 |
| L_caudalanteriorcingulate_thickavg | 0.010 | 0.469 | 18 |
| L_inferiorparietal_thickavg | 0.018 | 0.441 | 19 |
| L_entorhinal_thickavg | 0.022 | 0.426 | 20 |
| R_fusiform_thickavg | 0.024 | 0.420 | 21 |
| R_isthmuscingulate_thickavg | 0.029 | 0.419 | 22 |
| L_parsorbitalis_thickavg | 0.049 | 0.363 | 23 |
| L_cuneus_thickavg | 0.053 | 0.384 | 24 |
| L_inferiortemporal_thickavg | 0.054 | 0.358 | 25 |
| R_precuneus_thickavg | 0.071 | 0.314 | 26 |
| R_superiortemporal_thickavg | 0.076 | 0.289 | 27 |
| R_middletemporal_thickavg | 0.078 | 0.248 | 28 |
| R_caudalanteriorcingulate_thickavg | 0.081 | 0.260 | 29 |
| L_precuneus_thickavg | 0.088 | 0.255 | 30 |
| R_parahippocampal_thickavg | 0.088 | 0.237 | 31 |
| L_fusiform_thickavg | 0.106 | 0.195 | 32 |
| L_superiortemporal_thickavg | 0.110 | 0.194 | 33 |
| R_rostralmiddlefrontal_thickavg | 0.118 | 0.196 | 34 |
| R_pericalcarine_thickavg | 0.118 | 0.178 | 35 |
| L_postcentral_thickavg | 0.119 | 0.187 | 36 |
| L_superiorfrontal_thickavg | 0.121 | 0.164 | 37 |
| R_lateraloccipital_thickavg | 0.122 | 0.157 | 38 |
| L_supramarginal_thickavg | 0.129 | 0.142 | 39 |
| R_inferiorparietal_thickavg | 0.135 | 0.125 | 40 |
| L_parsopercularis_thickavg | 0.137 | 0.137 | 41 |
| R_supramarginal_thickavg | 0.150 | 0.103 | 42 |
| L_lateraloccipital_thickavg | 0.151 | 0.120 | 43 |
| R_bankssts_thickavg | 0.152 | 0.114 | 44 |
| R_lingual_thickavg | 0.155 | 0.135 | 45 |
| R_parsopercularis_thickavg | 0.156 | 0.135 | 46 |
| L_pericalcarine_thickavg | 0.158 | 0.102 | 47 |
| R_postcentral_thickavg | 0.166 | 0.111 | 48 |
| R_temporalpole_thickavg | 0.166 | 0.115 | 49 |
| L_superiorparietal_thickavg | 0.166 | 0.096 | 50 |
| R_superiorparietal_thickavg | 0.173 | 0.074 | 51 |
| R_posteriorcingulate_thickavg | 0.183 | 0.080 | 52 |
| L_temporalpole_thickavg | 0.183 | 0.079 | 53 |
| L_caudalmiddlefrontal_thickavg | 0.186 | 0.076 | 54 |
| L_lingual_thickavg | 0.188 | 0.098 | 55 |
| L_parstriangularis_thickavg | 0.191 | 0.069 | 56 |
| L_middletemporal_thickavg | 0.194 | 0.062 | 57 |
| L_bankssts_thickavg | 0.194 | 0.049 | 58 |
| R_cuneus_thickavg | 0.207 | 0.064 | 59 |
| L_rostralmiddlefrontal_thickavg | 0.217 | 0.038 | 60 |
| R_parstriangularis_thickavg | 0.236 | 0.037 | 61 |
| R_parsorbitalis_thickavg | 0.248 | 0.024 | 62 |
| R_precentral_thickavg | 0.257 | 0.037 | 63 |
| R_superiorfrontal_thickavg | 0.263 | 0.014 | 64 |
| L_paracentral_thickavg | 0.266 | 0.024 | 65 |
| L_precentral_thickavg | 0.278 | 0.017 | 66 |
| R_paracentral_thickavg | 0.286 | 0.018 | 67 |
| R_caudalmiddlefrontal_thickavg | 0.341 | 0.004 | 68 |

Table S10. Functional epicenters of amotivation-related cortical alteration pattern

| DKT_Region | r_value | pspin | Rank |
| --- | --- | --- | --- |
| R_rostralanteriorcingulate_thickavg | -0.295 | 0.009 | 1 |
| R_posteriorcingulate_thickavg | -0.243 | 0.025 | 2 |
| L_rostralanteriorcingulate_thickavg | -0.232 | 0.024 | 3 |
| L_transversetemporal_thickavg | -0.193 | 0.046 | 4 |
| L_caudalanteriorcingulate_thickavg | -0.185 | 0.057 | 5 |
| R_caudalanteriorcingulate_thickavg | -0.180 | 0.065 | 6 |
| L_posteriorcingulate_thickavg | -0.151 | 0.082 | 7 |
| R_supramarginal_thickavg | -0.124 | 0.154 | 8 |
| L_lateralorbitofrontal_thickavg | -0.117 | 0.089 | 9 |
| R_transversetemporal_thickavg | -0.113 | 0.165 | 10 |
| R_medialorbitofrontal_thickavg | -0.107 | 0.173 | 11 |
| R_superiortemporal_thickavg | -0.104 | 0.171 | 12 |
| R_precentral_thickavg | -0.103 | 0.170 | 13 |
| R_superiorfrontal_thickavg | -0.100 | 0.212 | 14 |
| L_insula_thickavg | -0.095 | 0.216 | 15 |
| R_parstriangularis_thickavg | -0.090 | 0.256 | 16 |
| L_postcentral_thickavg | -0.080 | 0.234 | 17 |
| L_isthmuscingulate_thickavg | -0.079 | 0.232 | 18 |
| L_parsopercularis_thickavg | -0.075 | 0.184 | 19 |
| L_paracentral_thickavg | -0.070 | 0.279 | 20 |
| L_superiortemporal_thickavg | -0.068 | 0.259 | 21 |
| R_temporalpole_thickavg | -0.067 | 0.297 | 22 |
| L_parahippocampal_thickavg | -0.066 | 0.266 | 23 |
| R_insula_thickavg | -0.065 | 0.302 | 24 |
| R_paracentral_thickavg | -0.063 | 0.262 | 25 |
| L_pericalcarine_thickavg | -0.062 | 0.293 | 26 |
| R_isthmuscingulate_thickavg | -0.055 | 0.301 | 27 |
| R_postcentral_thickavg | -0.053 | 0.302 | 28 |
| R_precuneus_thickavg | -0.051 | 0.329 | 29 |
| L_precentral_thickavg | -0.051 | 0.321 | 30 |
| L_precuneus_thickavg | -0.042 | 0.336 | 31 |
| R_rostralmiddlefrontal_thickavg | -0.040 | 0.433 | 32 |
| R_parahippocampal_thickavg | -0.035 | 0.357 | 33 |
| L_frontalpole_thickavg | -0.032 | 0.308 | 34 |
| R_pericalcarine_thickavg | -0.025 | 0.360 | 35 |
| R_middletemporal_thickavg | -0.024 | 0.421 | 36 |
| L_superiorparietal_thickavg | -0.023 | 0.448 | 37 |
| R_cuneus_thickavg | -0.012 | 0.442 | 38 |
| L_superiorfrontal_thickavg | -0.005 | 0.384 | 39 |
| L_rostralmiddlefrontal_thickavg | -0.003 | 0.443 | 40 |
| R_superiorparietal_thickavg | -0.001 | 0.482 | 41 |
| R_caudalmiddlefrontal_thickavg | 0.000 | 0.467 | 42 |
| L_lingual_thickavg | 0.001 | 0.503 | 43 |
| L_lateraloccipital_thickavg | 0.008 | 0.460 | 44 |
| R_lingual_thickavg | 0.009 | 0.485 | 45 |
| R_lateralorbitofrontal_thickavg | 0.011 | 0.378 | 46 |
| L_bankssts_thickavg | 0.014 | 0.545 | 47 |
| R_frontalpole_thickavg | 0.018 | 0.445 | 48 |
| L_medialorbitofrontal_thickavg | 0.018 | 0.519 | 49 |
| R_parsopercularis_thickavg | 0.020 | 0.397 | 50 |
| L_cuneus_thickavg | 0.029 | 0.382 | 51 |
| R_bankssts_thickavg | 0.030 | 0.411 | 52 |
| L_parstriangularis_thickavg | 0.032 | 0.494 | 53 |
| R_fusiform_thickavg | 0.038 | 0.381 | 54 |
| R_entorhinal_thickavg | 0.039 | 0.335 | 55 |
| R_parsorbitalis_thickavg | 0.043 | 0.350 | 56 |
| R_lateraloccipital_thickavg | 0.051 | 0.378 | 57 |
| L_caudalmiddlefrontal_thickavg | 0.057 | 0.422 | 58 |
| L_fusiform_thickavg | 0.060 | 0.304 | 59 |
| L_parsorbitalis_thickavg | 0.062 | 0.392 | 60 |
| L_temporalpole_thickavg | 0.069 | 0.380 | 61 |
| L_entorhinal_thickavg | 0.070 | 0.375 | 62 |
| R_inferiorparietal_thickavg | 0.073 | 0.246 | 63 |
| L_inferiorparietal_thickavg | 0.075 | 0.319 | 64 |
| R_inferiortemporal_thickavg | 0.080 | 0.229 | 65 |
| L_inferiortemporal_thickavg | 0.083 | 0.293 | 66 |
| L_supramarginal_thickavg | 0.086 | 0.242 | 67 |
| L_middletemporal_thickavg | 0.189 | 0.078 | 68 |

Table S11. Structural epicenters of amotivation-related cortical alteration pattern

| DKT_Region | r_value | pspin | Rank |
| --- | --- | --- | --- |
| R_rostralanteriorcingulate_thickavg | -0.410 | 0.001 | 1 |
| R_frontalpole_thickavg | -0.358 | 0.002 | 2 |
| R_caudalanteriorcingulate_thickavg | -0.348 | 0.006 | 3 |
| L_frontalpole_thickavg | -0.347 | 0.011 | 4 |
| R_superiorfrontal_thickavg | -0.313 | 0.005 | 5 |
| R_medialorbitofrontal_thickavg | -0.275 | 0.018 | 6 |
| L_rostralanteriorcingulate_thickavg | -0.274 | 0.002 | 7 |
| R_parsorbitalis_thickavg | -0.265 | 0.043 | 8 |
| L_lateralorbitofrontal_thickavg | -0.251 | 0.003 | 9 |
| L_caudalanteriorcingulate_thickavg | -0.219 | 0.014 | 10 |
| R_temporalpole_thickavg | -0.211 | 0.101 | 11 |
| R_posteriorcingulate_thickavg | -0.200 | 0.057 | 12 |
| R_parstriangularis_thickavg | -0.199 | 0.112 | 13 |
| R_lateralorbitofrontal_thickavg | -0.188 | 0.109 | 14 |
| R_rostralmiddlefrontal_thickavg | -0.171 | 0.121 | 15 |
| L_superiorfrontal_thickavg | -0.167 | 0.051 | 16 |
| L_posteriorcingulate_thickavg | -0.158 | 0.068 | 17 |
| R_precentral_thickavg | -0.123 | 0.224 | 18 |
| L_parsorbitalis_thickavg | -0.122 | 0.040 | 19 |
| R_insula_thickavg | -0.121 | 0.372 | 20 |
| R_caudalmiddlefrontal_thickavg | -0.116 | 0.219 | 21 |
| L_medialorbitofrontal_thickavg | -0.111 | 0.117 | 22 |
| R_parsopercularis_thickavg | -0.106 | 0.287 | 23 |
| R_superiorparietal_thickavg | -0.073 | 0.379 | 24 |
| R_postcentral_thickavg | -0.061 | 0.351 | 25 |
| R_isthmuscingulate_thickavg | -0.058 | 0.435 | 26 |
| L_precuneus_thickavg | -0.057 | 0.200 | 27 |
| R_pericalcarine_thickavg | -0.054 | 0.378 | 28 |
| R_middletemporal_thickavg | -0.054 | 0.555 | 29 |
| R_precuneus_thickavg | -0.052 | 0.460 | 30 |
| L_bankssts_thickavg | -0.035 | 0.237 | 31 |
| R_supramarginal_thickavg | -0.033 | 0.524 | 32 |
| L_transversetemporal_thickavg | -0.033 | 0.241 | 33 |
| L_temporalpole_thickavg | -0.011 | 0.259 | 34 |
| R_paracentral_thickavg | -0.010 | 0.430 | 35 |
| R_parahippocampal_thickavg | -0.009 | 0.676 | 36 |
| L_caudalmiddlefrontal_thickavg | -0.005 | 0.373 | 37 |
| R_transversetemporal_thickavg | -0.003 | 0.561 | 38 |
| L_rostralmiddlefrontal_thickavg | 0.000 | 0.630 | 39 |
| R_superiortemporal_thickavg | 0.010 | 0.287 | 40 |
| L_postcentral_thickavg | 0.011 | 0.538 | 41 |
| L_parsopercularis_thickavg | 0.014 | 0.650 | 42 |
| L_lingual_thickavg | 0.019 | 0.616 | 43 |
| L_parstriangularis_thickavg | 0.022 | 0.595 | 44 |
| R_lingual_thickavg | 0.023 | 0.290 | 45 |
| L_paracentral_thickavg | 0.030 | 0.417 | 46 |
| L_insula_thickavg | 0.030 | 0.771 | 47 |
| L_superiorparietal_thickavg | 0.033 | 0.547 | 48 |
| L_isthmuscingulate_thickavg | 0.039 | 0.468 | 49 |
| R_fusiform_thickavg | 0.046 | 0.183 | 50 |
| R_inferiorparietal_thickavg | 0.047 | 0.166 | 51 |
| L_precentral_thickavg | 0.052 | 0.438 | 52 |
| R_cuneus_thickavg | 0.058 | 0.266 | 53 |
| L_parahippocampal_thickavg | 0.060 | 0.526 | 54 |
| L_inferiorparietal_thickavg | 0.075 | 0.421 | 55 |
| R_lateraloccipital_thickavg | 0.075 | 0.150 | 56 |
| L_pericalcarine_thickavg | 0.084 | 0.310 | 57 |
| L_cuneus_thickavg | 0.087 | 0.271 | 58 |
| R_entorhinal_thickavg | 0.088 | 0.131 | 59 |
| L_entorhinal_thickavg | 0.104 | 0.275 | 60 |
| L_superiortemporal_thickavg | 0.113 | 0.320 | 61 |
| L_inferiortemporal_thickavg | 0.136 | 0.208 | 62 |
| R_inferiortemporal_thickavg | 0.140 | 0.048 | 63 |
| R_bankssts_thickavg | 0.160 | 0.047 | 64 |
| L_supramarginal_thickavg | 0.183 | 0.091 | 65 |
| L_lateraloccipital_thickavg | 0.218 | 0.048 | 66 |
| L_fusiform_thickavg | 0.221 | 0.040 | 67 |
| L_middletemporal_thickavg | 0.222 | 0.045 | 68 |

Table S12. Functional epicenters of diminished expression-related cortical alteration pattern

| DKT_Region | r_value | pspin | Rank |
| --- | --- | --- | --- |
| R_rostralanteriorcingulate_thickavg | -0.172 | 0.096 | 1 |
| R_temporalpole_thickavg | -0.118 | 0.184 | 2 |
| L_rostralanteriorcingulate_thickavg | -0.103 | 0.211 | 3 |
| L_medialorbitofrontal_thickavg | -0.093 | 0.225 | 4 |
| R_superiortemporal_thickavg | -0.082 | 0.252 | 5 |
| R_medialorbitofrontal_thickavg | -0.072 | 0.258 | 6 |
| R_posteriorcingulate_thickavg | -0.070 | 0.285 | 7 |
| L_insula_thickavg | -0.042 | 0.342 | 8 |
| L_parstriangularis_thickavg | -0.034 | 0.385 | 9 |
| R_precentral_thickavg | -0.033 | 0.377 | 10 |
| L_transversetemporal_thickavg | -0.027 | 0.414 | 11 |
| L_parahippocampal_thickavg | -0.025 | 0.408 | 12 |
| R_frontalpole_thickavg | -0.024 | 0.433 | 13 |
| L_entorhinal_thickavg | -0.022 | 0.413 | 14 |
| L_temporalpole_thickavg | -0.016 | 0.474 | 15 |
| L_lateraloccipital_thickavg | -0.016 | 0.425 | 16 |
| L_isthmuscingulate_thickavg | -0.014 | 0.420 | 17 |
| R_caudalanteriorcingulate_thickavg | -0.008 | 0.468 | 18 |
| R_superiorparietal_thickavg | -0.007 | 0.483 | 19 |
| L_precuneus_thickavg | -0.005 | 0.465 | 20 |
| R_parsorbitalis_thickavg | -0.003 | 0.465 | 21 |
| L_posteriorcingulate_thickavg | -0.001 | 0.480 | 22 |
| L_fusiform_thickavg | 0.001 | 0.520 | 23 |
| R_isthmuscingulate_thickavg | 0.003 | 0.501 | 24 |
| R_superiorfrontal_thickavg | 0.015 | 0.473 | 25 |
| R_parahippocampal_thickavg | 0.017 | 0.458 | 26 |
| R_supramarginal_thickavg | 0.018 | 0.459 | 27 |
| R_paracentral_thickavg | 0.018 | 0.455 | 28 |
| L_caudalmiddlefrontal_thickavg | 0.023 | 0.420 | 29 |
| L_superiortemporal_thickavg | 0.024 | 0.449 | 30 |
| R_fusiform_thickavg | 0.025 | 0.440 | 31 |
| R_rostralmiddlefrontal_thickavg | 0.026 | 0.424 | 32 |
| L_postcentral_thickavg | 0.027 | 0.468 | 33 |
| R_parstriangularis_thickavg | 0.027 | 0.414 | 34 |
| R_precuneus_thickavg | 0.028 | 0.411 | 35 |
| L_precentral_thickavg | 0.029 | 0.432 | 36 |
| R_insula_thickavg | 0.031 | 0.425 | 37 |
| L_caudalanteriorcingulate_thickavg | 0.033 | 0.415 | 38 |
| L_superiorfrontal_thickavg | 0.038 | 0.349 | 39 |
| L_parsopercularis_thickavg | 0.040 | 0.348 | 40 |
| L_paracentral_thickavg | 0.042 | 0.401 | 41 |
| L_inferiortemporal_thickavg | 0.045 | 0.314 | 42 |
| R_bankssts_thickavg | 0.049 | 0.356 | 43 |
| L_pericalcarine_thickavg | 0.049 | 0.369 | 44 |
| L_frontalpole_thickavg | 0.051 | 0.310 | 45 |
| R_postcentral_thickavg | 0.056 | 0.322 | 46 |
| R_entorhinal_thickavg | 0.057 | 0.345 | 47 |
| L_supramarginal_thickavg | 0.063 | 0.342 | 48 |
| R_inferiorparietal_thickavg | 0.065 | 0.301 | 49 |
| L_cuneus_thickavg | 0.066 | 0.326 | 50 |
| R_cuneus_thickavg | 0.077 | 0.287 | 51 |
| L_superiorparietal_thickavg | 0.078 | 0.302 | 52 |
| R_middletemporal_thickavg | 0.083 | 0.259 | 53 |
| L_bankssts_thickavg | 0.087 | 0.252 | 54 |
| L_rostralmiddlefrontal_thickavg | 0.088 | 0.223 | 55 |
| R_transversetemporal_thickavg | 0.092 | 0.246 | 56 |
| L_parsorbitalis_thickavg | 0.099 | 0.188 | 57 |
| R_lateraloccipital_thickavg | 0.100 | 0.205 | 58 |
| L_inferiorparietal_thickavg | 0.102 | 0.209 | 59 |
| R_pericalcarine_thickavg | 0.108 | 0.200 | 60 |
| R_lateralorbitofrontal_thickavg | 0.112 | 0.206 | 61 |
| L_lateralorbitofrontal_thickavg | 0.116 | 0.172 | 62 |
| R_lingual_thickavg | 0.121 | 0.177 | 63 |
| L_middletemporal_thickavg | 0.126 | 0.129 | 64 |
| R_caudalmiddlefrontal_thickavg | 0.133 | 0.158 | 65 |
| L_lingual_thickavg | 0.135 | 0.162 | 66 |
| R_parsopercularis_thickavg | 0.161 | 0.102 | 67 |
| R_inferiortemporal_thickavg | 0.205 | 0.054 | 68 |

Table S12. Structural epicenters of diminished expression-related cortical alteration pattern

| DKT_Region | r_value | pspin |
| --- | --- | --- |
| R_temporalpole_thickavg | -0.348 | 0.005 |
| L_medialorbitofrontal_thickavg | -0.217 | 0.045 |
| R_frontalpole_thickavg | -0.178 | 0.080 |
| L_rostralanteriorcingulate_thickavg | -0.177 | 0.067 |
| R_rostralanteriorcingulate_thickavg | -0.151 | 0.131 |
| R_caudalanteriorcingulate_thickavg | -0.133 | 0.157 |
| L_bankssts_thickavg | -0.122 | 0.124 |
| R_parsorbitalis_thickavg | -0.117 | 0.167 |
| R_fusiform_thickavg | -0.103 | 0.229 |
| L_caudalanteriorcingulate_thickavg | -0.094 | 0.188 |
| R_superiortemporal_thickavg | -0.090 | 0.240 |
| R_parahippocampal_thickavg | -0.075 | 0.265 |
| R_medialorbitofrontal_thickavg | -0.065 | 0.278 |
| L_frontalpole_thickavg | -0.060 | 0.306 |
| L_posteriorcingulate_thickavg | -0.057 | 0.346 |
| R_superiorparietal_thickavg | -0.051 | 0.302 |
| R_bankssts_thickavg | -0.041 | 0.324 |
| R_superiorfrontal_thickavg | -0.038 | 0.374 |
| R_supramarginal_thickavg | -0.038 | 0.361 |
| R_inferiorparietal_thickavg | -0.026 | 0.412 |
| L_lateralorbitofrontal_thickavg | -0.024 | 0.456 |
| R_lingual_thickavg | -0.023 | 0.414 |
| R_insula_thickavg | -0.022 | 0.356 |
| R_parstriangularis_thickavg | -0.019 | 0.429 |
| R_posteriorcingulate_thickavg | -0.010 | 0.427 |
| L_parstriangularis_thickavg | -0.003 | 0.511 |
| R_entorhinal_thickavg | -0.002 | 0.480 |
| L_temporalpole_thickavg | 0.000 | 0.474 |
| R_cuneus_thickavg | 0.009 | 0.494 |
| L_superiorfrontal_thickavg | 0.014 | 0.472 |
| R_lateralorbitofrontal_thickavg | 0.022 | 0.437 |
| L_transversetemporal_thickavg | 0.022 | 0.412 |
| R_middletemporal_thickavg | 0.024 | 0.448 |
| R_transversetemporal_thickavg | 0.027 | 0.458 |
| L_precuneus_thickavg | 0.031 | 0.393 |
| R_isthmuscingulate_thickavg | 0.036 | 0.389 |
| R_pericalcarine_thickavg | 0.040 | 0.390 |
| R_inferiortemporal_thickavg | 0.041 | 0.396 |
| L_parsorbitalis_thickavg | 0.042 | 0.314 |
| L_lateraloccipital_thickavg | 0.043 | 0.313 |
| L_caudalmiddlefrontal_thickavg | 0.044 | 0.350 |
| L_entorhinal_thickavg | 0.047 | 0.300 |
| R_rostralmiddlefrontal_thickavg | 0.047 | 0.364 |
| L_parahippocampal_thickavg | 0.057 | 0.234 |
| R_precuneus_thickavg | 0.062 | 0.327 |
| R_caudalmiddlefrontal_thickavg | 0.063 | 0.346 |
| L_insula_thickavg | 0.063 | 0.185 |
| L_supramarginal_thickavg | 0.065 | 0.219 |
| L_parsopercularis_thickavg | 0.070 | 0.209 |
| R_precentral_thickavg | 0.070 | 0.326 |
| L_inferiorparietal_thickavg | 0.071 | 0.232 |
| L_isthmuscingulate_thickavg | 0.075 | 0.288 |
| L_middletemporal_thickavg | 0.086 | 0.150 |
| R_postcentral_thickavg | 0.090 | 0.268 |
| L_lingual_thickavg | 0.092 | 0.190 |
| L_superiortemporal_thickavg | 0.094 | 0.136 |
| L_superiorparietal_thickavg | 0.102 | 0.189 |
| R_lateraloccipital_thickavg | 0.104 | 0.226 |
| L_cuneus_thickavg | 0.106 | 0.198 |
| L_paracentral_thickavg | 0.108 | 0.190 |
| L_fusiform_thickavg | 0.115 | 0.116 |
| R_paracentral_thickavg | 0.130 | 0.169 |
| L_postcentral_thickavg | 0.133 | 0.139 |
| L_pericalcarine_thickavg | 0.151 | 0.091 |
| L_inferiortemporal_thickavg | 0.177 | 0.038 |
| L_rostralmiddlefrontal_thickavg | 0.188 | 0.059 |
| L_precentral_thickavg | 0.195 | 0.040 |
| R_parsopercularis_thickavg | 0.208 | 0.069 |
